# MedTech innovation identification: A rapid scoping review of patent research studies to inform horizon scanning methods

**DOI:** 10.1101/2024.12.09.24318714

**Authors:** Sonia Garcia Gonzalez-Moral, Erin Pennock, Olushola Ewedairo, Elizabeth Green, James Elgey, Andrew Mkwashi

## Abstract

**Objectives:** This study investigates the role of and methods for patent analysis in advancing medical technology (MedTech) innovation, a sector characterized by diverse, non-pharmacological or non-immunological healthcare technologies and significant research investment. Patents are critical early indicators of innovation, supporting horizon scanning and weak signal detection. The study aimed to identify intellectual property sources, evaluate methods for patent retrieval and analysis, and outline objectives for using patent data to anticipate trends and inform healthcare strategies. It also offered a methodological framework to support stakeholders in adopting innovative MedTech solutions.

**Methods:** A rapid review (RR) was conducted using Cochrane Rapid Review Methods and PRISMA guidelines, with a pre-registered protocol on the Open Science Framework. Searches in Embase, IEEE Xplore, and Web of Science targeted records from 2020 onwards. Three independent reviewers screened studies using Rayyan. We included any study type, published since 2020 that provided sufficient data on patent sources, methods and tools applied to the study of MedTech. Our data extraction included bibliographic details, study characteristics, and methodological information. Risk of bias assessments were not undertaken. Narrative and tabular methods, supplemented by visual charts, were employed to synthesise findings.

**Results:** Our searches identified 1,741 studies, of which 124 were included after title, abstract, and full-text screening, with 54% being original research, 44% reviews, and the remainder being conference abstracts. Most studies (68%) relied solely on patent databases, while others searched the grey literature. Research objectives of the included studies were grouped into nine themes, with trend analysis (50%) and policy recommendations (20%) being the most common. The review analysed 199 patent databases, with 27% of studies using multiple sources. Time horizons for patent searches averaged 24.6 years, ranging from 1900 to 2019. Automated approaches, employed in 33% of studies, frequently utilised tools like Gephi for network visualization. Disease mapping, based on NICE classification, indicated that cancer (19%) and respiratory conditions (16%), particularly COVID-19, were key areas, while digital health dominated the "health and social care delivery" category.

**Conclusions:** The review highlights the value of patent data in trend analysis and its broader role in shaping policies and research strategies. While patents provide crucial insights into emerging technologies, inconsistent de-duplication practices across studies pose a risk of data inflation, accentuating the need for transparency and rigour. Finally, this review emphasized the importance of data transformation and visualization in detecting emerging trends with Python and R being the most commonly used programming languages for developing custom tools.

## Introduction

Medical devices and invitro diagnostics, henceforth MedTech, are a heterogeneous group of healthcare technologies that are mainly characterised by the fact that they do not achieve their primary intended action by pharmacological, immunological or metabolic means.[1] Several definitions exist, with the UK Medical Device Regulation (2002)[2] and the International Medical Device Regulators Forum[1] definitions coinciding in many of the aspects that characterise a medical device. Medical technology is defined by continuous innovation, with an estimated 8% of overall investment in the sector directed towards research and development. Circa 16,000 patent applications were filed in the European Patent Office (EPO) in 2023 in the field of medical technology.[3]

A thorough understanding of the MedTech innovation pathway from ideation to market is imperative for strategic decision-making in healthcare, the formulation of health policy, and the allocation of research and development funds. It can also play a vital role in anticipating regulatory challenges associated with emerging technologies, such as those triggered by the integration of software and artificial intelligence (AI) in MedTech. Additionally, this knowledge facilitates early awareness for MedTech guidance and guideline documents developers, health service providers, and other stakeholders involved in the advancement and implementation of innovative healthcare solutions.

According to the World Intellectual property Office (WIPO) “a patent is an exclusive right granted for an invention”.[4] From the moment a patent is granted, a full disclosure of the invention to a patent office would have occurred and this information would have been made available to the public.[4] In the MedTech context, this includes medical devices and diagnostics, where clinical trials are required for some but not all. Often inventors will apply for patents before clinical trials are initiated to protect the intellectual property of the innovative technology in the trial. However, in the case of low-risk medical devices, where clinical trials are not always conducted, patents may be one of the few indications of a new innovative product before it is introduced to the market. Therefore, patents may be considered as one of the first signs of innovation and are often searched in horizon scanning studies alone or in combination with other sources of weak signal detection.[5]

Examining patent data offers many advantages such as understanding a particular technology’s evolution, allowing us to assess an invention and anticipate future trends.[6] Patents also serve as useful indicators for gauging a firm’s level of innovation and can act as a proxy for identifying connections between innovators.[7][8] Detecting signs of innovation ahead of being available on the market is one of the main objectives of the Innovation Observatory, a national horizon scanning and research intelligence body funded by the National Institute for Health and Care Research and hosted by Newcastle University in the UK. This study has three primary aims: identifying sources of intellectual property, such as patent databases; exploring the various methods used for patent retrieval and analysis; and determining the key objectives for employing patent analyses in the context of MedTech innovation. The findings are intended to offer a methodological foundation for utilizing patent analysis in horizon scanning, particularly for technologies in their early stages of development.

## Methods

A rapid review (RR) to scope the published literature on methods for patent landscape analysis was undertaken in line with the recommendations published by the Cochrane Rapid Review Methods working group.[9] We followed the Preferred Reporting Items for Systematic Reviews and Meta-analyses (PRISMA) extension for scoping reviews (ScRs) for the reporting of items relevant to this rapid review due to the lack of PRISMA reporting standards for RRs. This review did not quality appraise the included studies as it was considered out of scope for the objectives of this project. A pre-agreed protocol outlining the methods and objectives of this review was registered in Open Science Framework in June 2024[10] and is available for consultation here: https://doi.org/10.31219/osf.io/jpruk

## Inclusion criteria

We followed the Cochrane guide for outlining the inclusion criteria of this methods RR. These followed the study type, data, methods and outcomes (SDMO) framework.[11] Studies eligible for inclusion needed to meet the following criteria: (1) any type of study or conference abstract that reported sufficient information on methods and sources in English language; (2) data related to the health technology were sufficiently reported; (3) methods and tools used for the analysis were mentioned and (4) heterogenous outcomes were considered for inclusion although not considered mandatory for the inclusion of studies in this review in line with the recommendation from Higgins et al.[11]

## Search strategy

Our search strategy consisted of subject heading terms in combination with text words. A targeted search strategy was devised and run in Embase (Ovid) by an experienced information specialist and checked against the PRESS checklist.[12] This search was then translated and run in two more databases selected for their comprehensive coverage of technological and scientific topics: IEEE Xplore digital library and Web of Science. We searched in the title, abstract and keyword fields. Time limits were imposed to retrieve records published since 2020 to capture the most current tools and sources. No language limit or any other limits were used. Records were downloaded into Endnote 20 (Clarivate Analytics) for de-duplication. Full search strategies with results are porovided in Appendix A.

## Data collection

De-duplicated results were screened at the title and abstract levels by three independent reviewers separately using Rayyan, a screening software for systematic reviews.[13] Rayyan has an in-built tool for prioritisation of relevant records based on reviewers ranking, this tool was not used in this instance due to the relatively small number of de-duplicated records and the reviewer capacity available for this project. Consultation between reviewers was practiced at this stage in multiple occasions, in case of doubt due to ambiguity of titles or lack of clear reporting in abstracts, records were included for full-text screening. In this stage the same reviewers proceeded independently to screen the full texts for inclusion. One-to-one consultation was exercised regularly throughout this process and disagreements were resolved by checking with a fourth reviewer.

## Data extraction

A data extraction form was devised in line with the agreed objectives of this review as reported in the protocol.[10] Data was extracted by three independent reviewers and quality assessed by one for cleaning and standardising data ahead of analysis and synthesis. We extracted bibliographic details, study-related characteristics such as the type of technologies, health condition and objectives of the study, methods-related data including the sources searched, the geographical coverage of the sources, the time period searched and the use, if any, of automated methods and tools.

## Data synthesis

We used narrative and tabulated methods for summarising and synthesizing data collected during our data extraction stage. As a result of using narrative methods, we were able to explore the data in more depth by providing context and uncovering underlying themes. By weaving together qualitative insights, narrative methods offered a more comprehensive understanding of our research findings. Whenever possible, charts have been used to visually present information relevant to the objectives of this study.

## Results

Our searches identified a total of 1,741 records. After de-duplication, 1,505 remained. We sought the full text of 261 records of which only 124 were considered for inclusion in this review. Figure 1 presents a PRISMA flowchart with the retrieval and selection process for this review.

**Figure 1:**
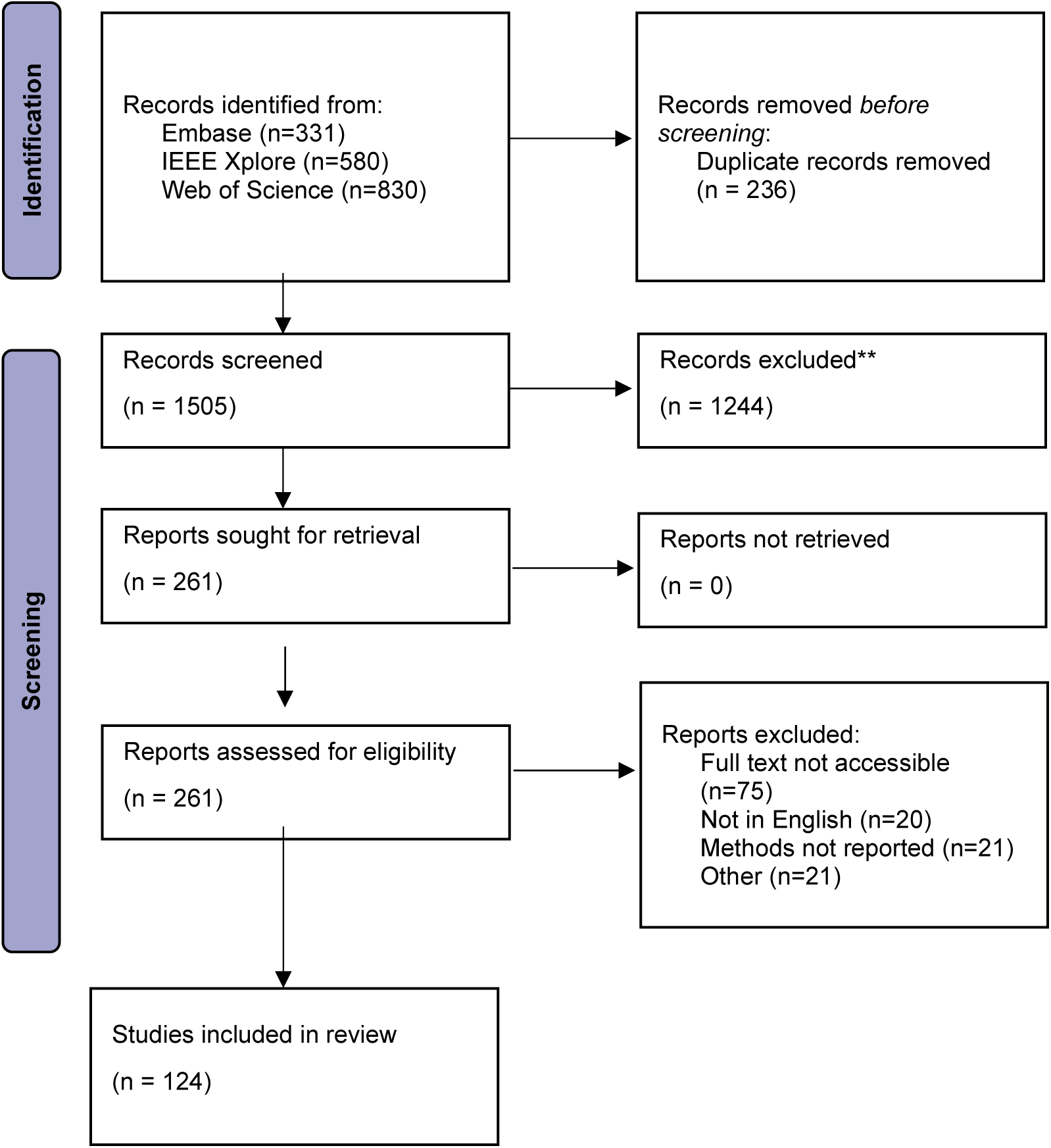
PRISMA flowchart for study selection

**Figure 2:**
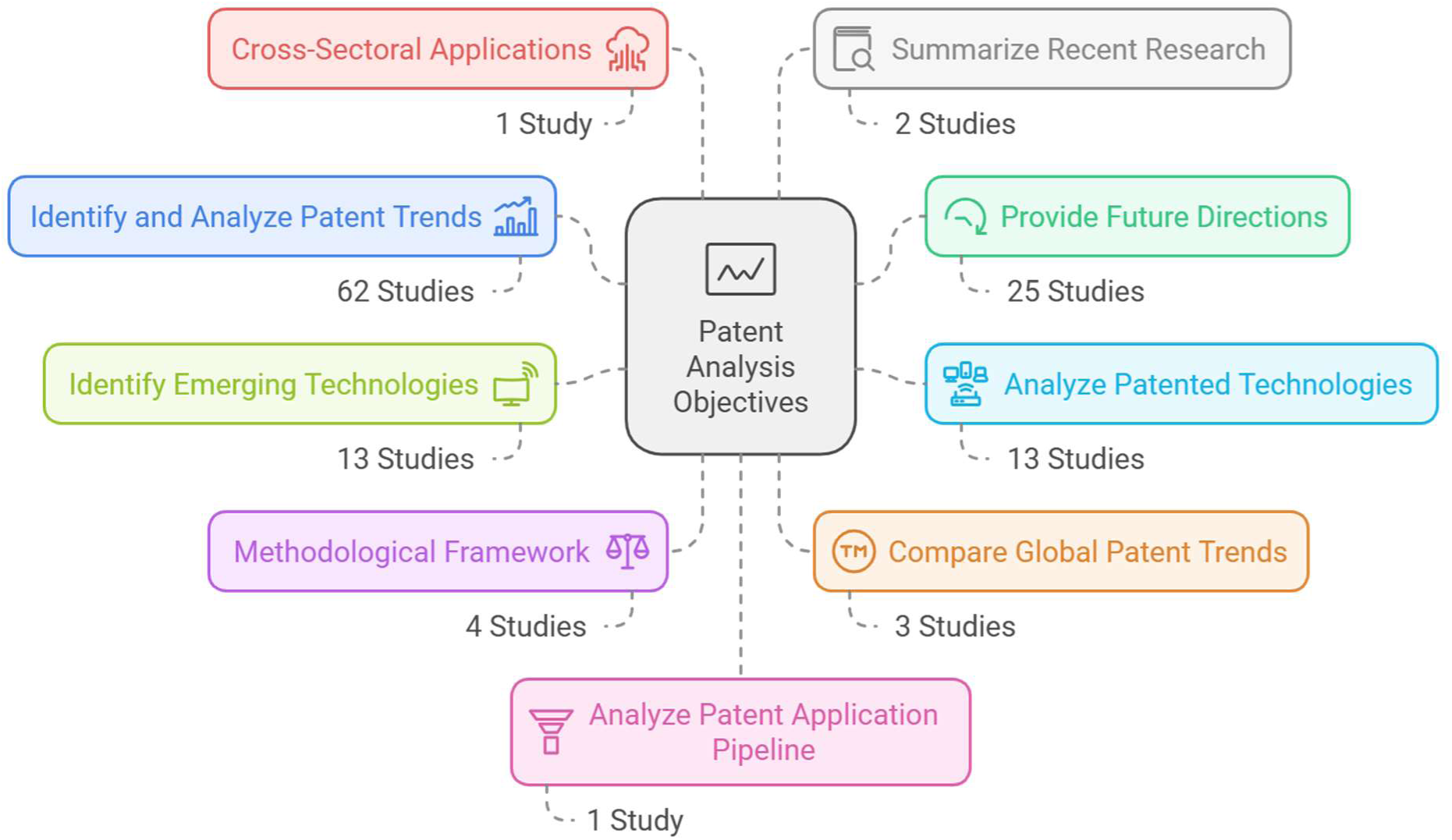
Reasons for conducting a patent research study by themes

## Study characteristics

From the 124 studies included, 67 (54%) were original research papers, 55 (44%) corresponded to reviews, 2 were conference papers and 1 short presentation. The majority of sources considered patent databases only (85, 68%) with the rest of included studies using a combination of different sources. These studies were published in 94 different journals. Nature Biotechnology was the most frequent journal in our dataset (n=8 studies). In 2020, there were 21 papers published, followed by a rise to 33 publications in 2021. The number of papers remained high but plateaued with 31 publications in both 2022 and 2023. By 2024, 8 papers had been published at the time of the review (July 2024). A table outlining the characteristics of the included studies is presented in Appendix B.

## Research objectives of included studies

The objectives of each paper were summarised and grouped into nine broad themes. The most common reason reported for the study of patents (62 studies, 50%) was to investigate trends within specific fields, followed by those aiming to provide recommendations for future research, policy, and strategy development (25 studies, 20%). Two other frequent objectives were: the study of specific patented technologies (13 studies, 10%) and the identification of emerging technologies (13 studies, 10%). Less frequent objectives included: the development of methodological frameworks for patent analysis (4 studies), analysis of global patent trends (3 studies), cross-sectoral applications of emerging technologies (1 study) and summary of recent research in specific fields (2 study).

## Sources of patent data

Eighty-five studies used patents alone, whereas 39 studies used various sources which included grey literature, research papers and clinical trials. Two studies did not report which patent database had been used. Over 27% of papers described the use of more than one patent database, as such across the 124 papers, there were 199 databases listed. Figure 3 presents the distribution of the top ten named databases across all papers.

**Figure 3:**
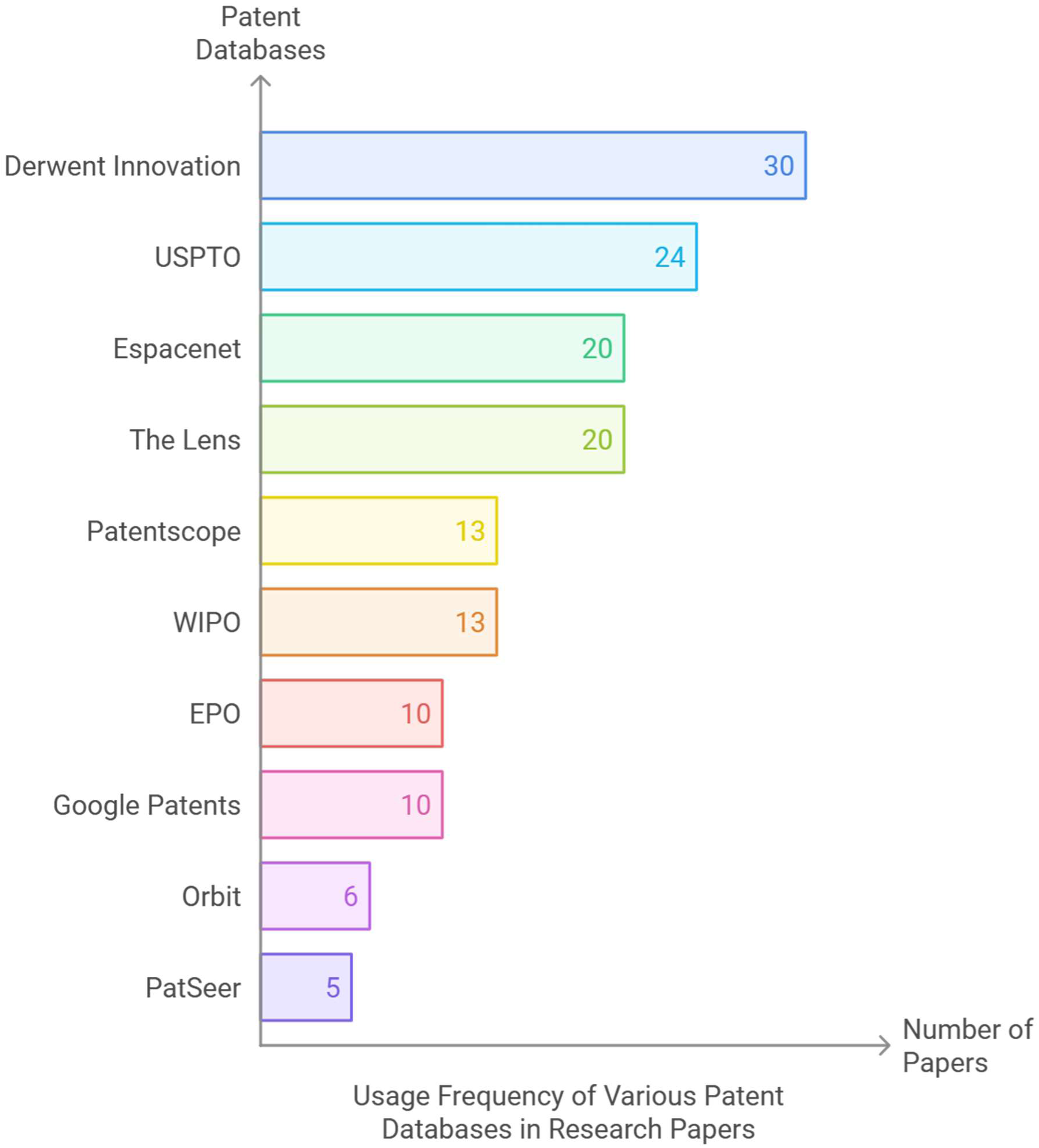
Top ten patent databases used in the included studies USPTO: United States Patent and Trademark Office; WIPO: World Intellectual Property Office; EPO: European Patent Office

**Figure 4.**
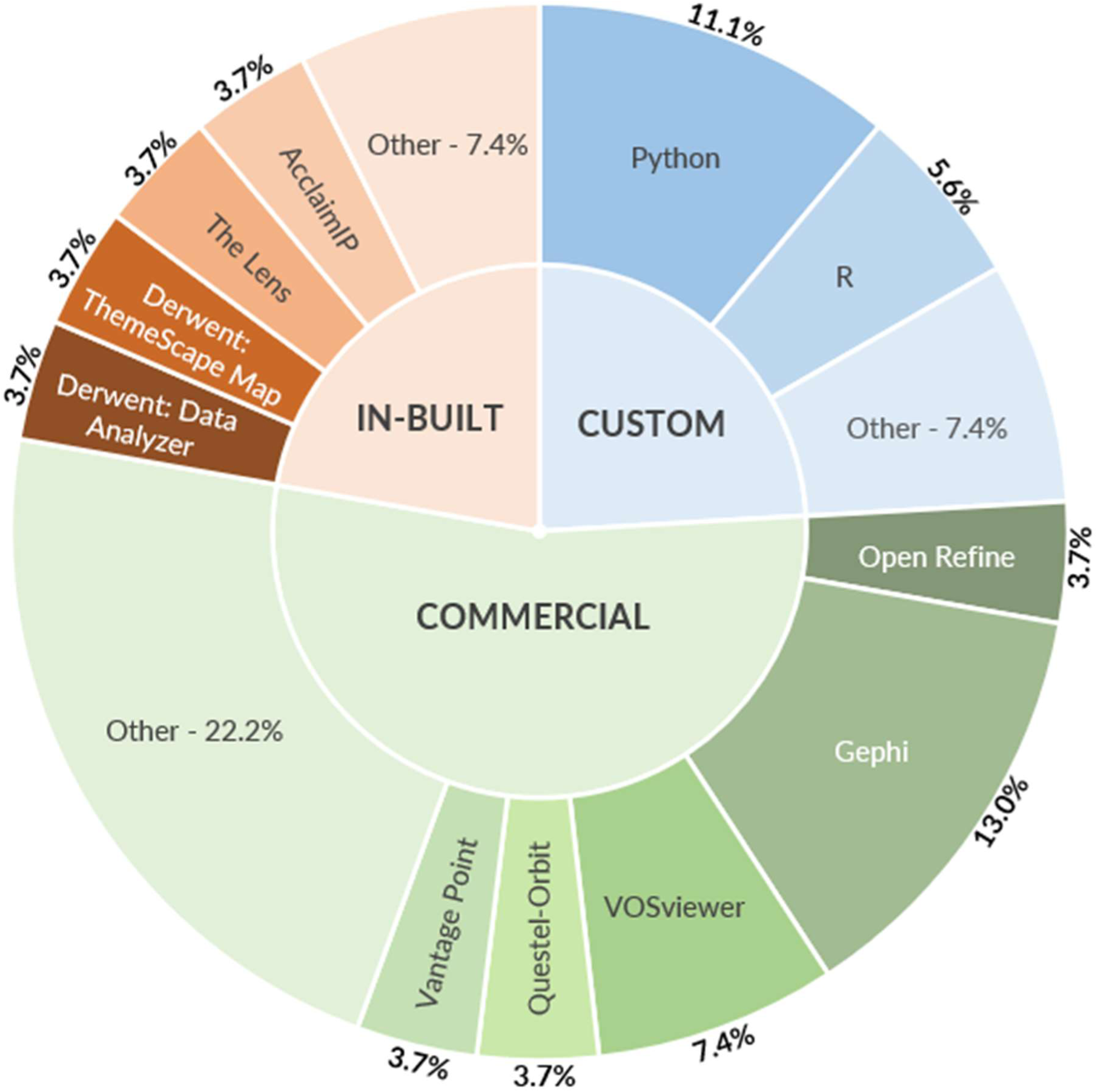
Reported tools used for automated or semi-automated approaches in this study.

**Figure 5.**
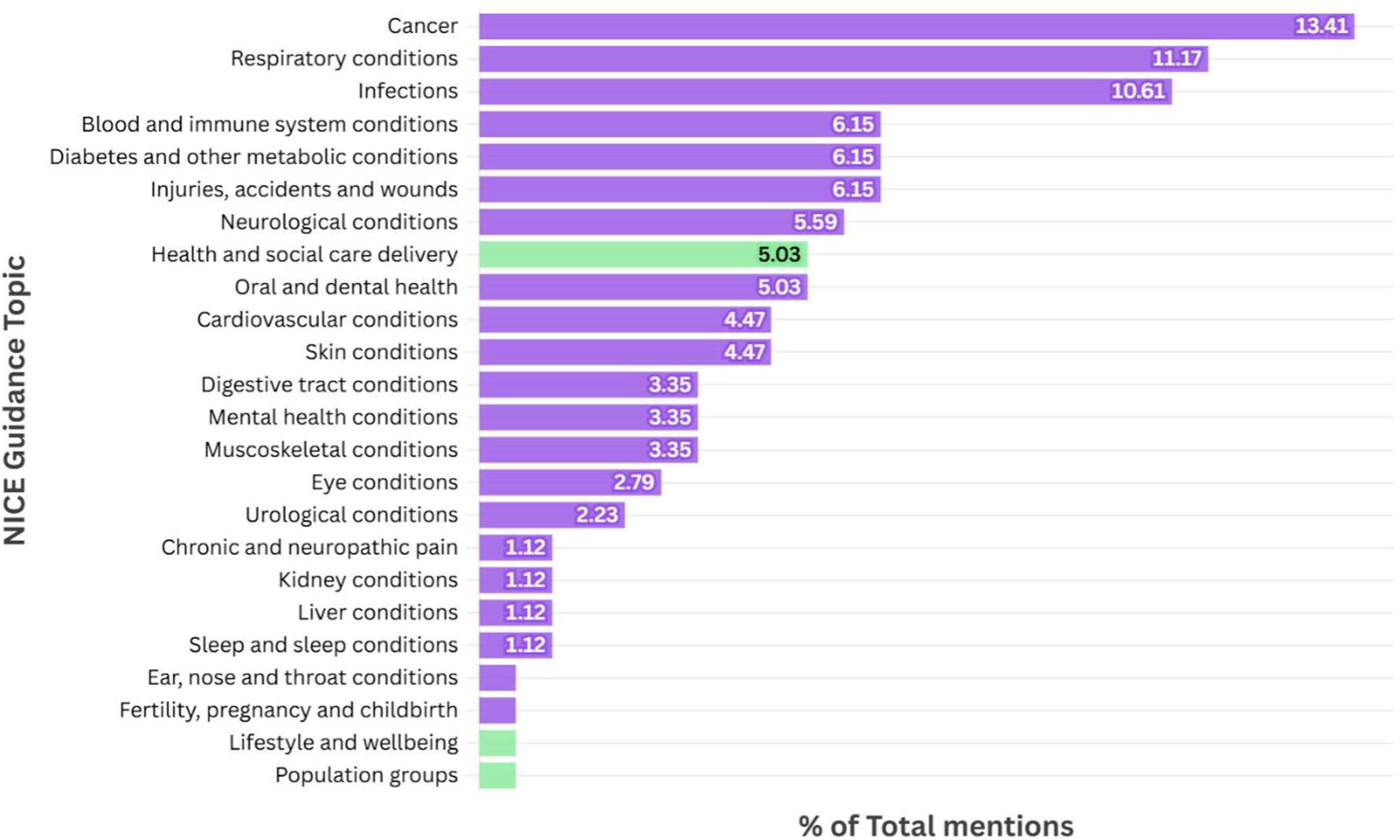
Specific mentions of NICE disease (purple) and other (green) guidance topics as a percentage of 178 total NICE guidance topics referenced in the results of 94 papers that reported specific NICE topics in their results.

## Time Horizons of Patent Scans

Of the 124 included papers 46 (37%) papers did not report the time period searched during patent analysis or gave incomplete timelines with only an end date leaving 78 papers with completed data on time horizons.

The mean length of time horizon for patent searches was 24.6 years. Only 4 papers searched a time period of over 50 years with the broadest time period searched being from 1900 to 2019.[127] Around 19% of papers with a reported time horizon searched a period of between 3 months and ten years. Up to 15% of papers searched a 20 year period. It was common for authors to backdate searches to start of the year 2000, although rationale was not provided.

## Methods and Automated Approaches Used

Of the 124 included papers 41 (32.8%) reported the use of automated or semi-automated approaches during patent analysis. Of these 41 papers, there were 62 explicit mentions of 43 unique tools that were used for patent analysis. Figure 3 presents all tools categorised into whether the tool was available as an in-built analysis module by the patent database searched (IN-BUILT), a custom patent analysis script (CUSTOM) or a commercially offered tool (COMMERCIAL). A full account of each tool used and their frequency across the 41 studies is available in Table 1.

**Table 1.**
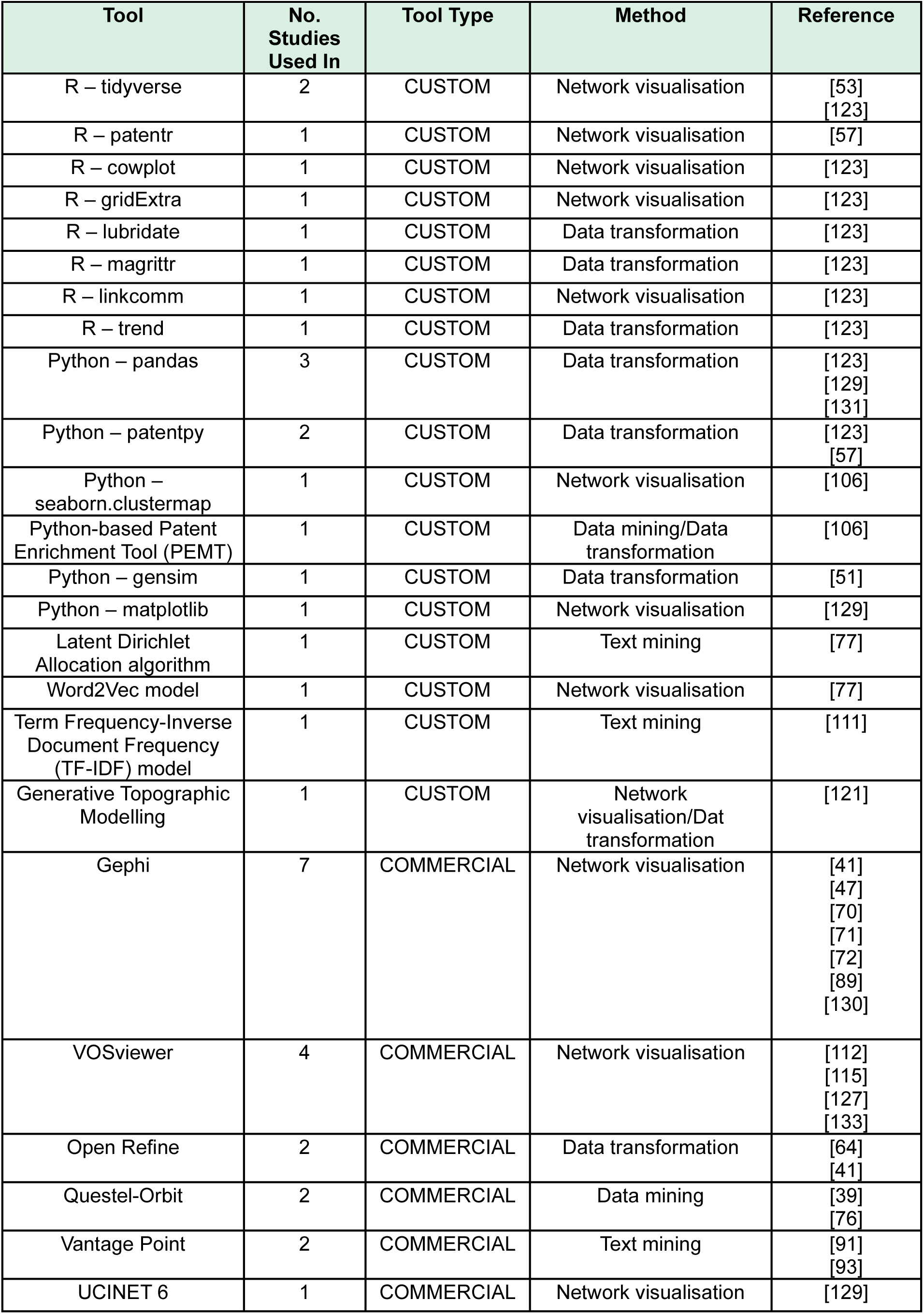
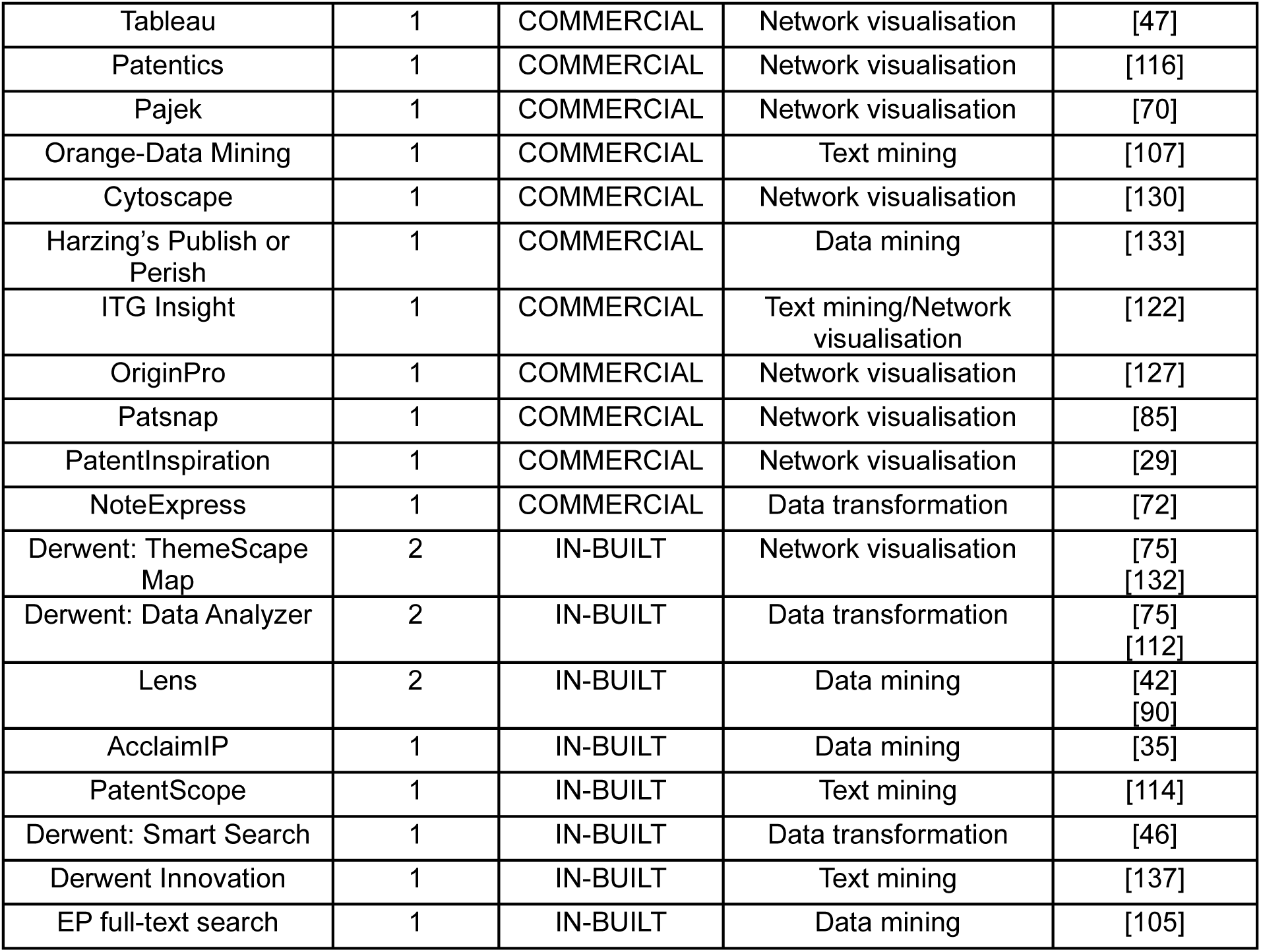
Full account of all named tools used for automated or semi-automated approaches for patent analysis from 41 papers which reported their use.

These tools were employed mainly in four different methods: data mining (understood as the statistical technique of processing raw data in a structured form), text mining (the part of data mining which involves processing of text from documents),[134] data transformation (the process of converting and cleaning raw data from one data source to meet the requirements of its new location)[135] and network visualisation (understood as the graphical representations of network devices, network metrics, and data flows).[136] Of the 64 methods discussed in this subsection, network visualisation was by far the most popular, used in 50% of the methods described. The most popular tool for network visualisation was the freely available software Gephi, used in 7 separate studies. Table 2 presents a breakdown of these tools classified by their type and the number of studies that used them.

### NICE Disease Classification Mapping

We used the National Institute for Health and Care Excellence (NICE) disease classification system[138] to map the 94 studies (75%) that reported specific disease indications or population groups. A total of 178 separate topic references throughout the 94 papers were extracted and mapped against the NICE classification. Despite some second level disease indications (e.g., COVID-19) that can be mapped to multiple NICE first level disease classifications, each mention of disease was only mapped one to avoid inflation of results.

Overall, ‘Cancer’ was the most prevalent NICE topic with 19% of the 124 included studies reporting cancer patents as either the primary or secondary finding. Of the 16% of papers that reported ‘Respiratory conditions’, 60% of these were specific to ‘COVID-19’ or other coronaviruses. ‘Health and social care delivery’ was the most popular NICE topic discussed in the included studies with 67% of these pertaining to ‘Digital health’.

## Discussion

In the MedTech context, the continuous search for signs of innovation is driving methods and tools development. This rapid scoping review has revealed an array of methodological approaches, tools and reasons why patents are being used to discover these signals. We used rapid review methods for the identification of these studies which usually involve a trade-off between sensitivity and specificity of the searches to allow processing search results in a rapid context. However, our high level of included studies indicates that precision of patent-related keywords may have positively contributed to increase the sensitivity of the search without increasing the recall.

Our review identified just under 200 different sources for patent data retrieval and some studies used more than one source. While the majority of the studies used patent databases, other studies extended their searches into clinical trials and published literature. Several studies have superficially provided descriptions of de-duplication processes used to manage the large quantities of patent data obtained when more than one database was used. However, failing to apply thorough de-duplication methods could lead to over estimation of patent data and innovation clusters. If de-duplication is used, this step should be undertaken before patent analyses to eliminate inflated patent data and represent more realistically the strength and size of the weak signals. If de-duplication is not undertaken, the rationale for that should also be transparently reported.

As a result of the abundance and easy accessibility of global patent data, patent data is mostly used in trend analysis, aligning with methods used for scanning emerging technologies and predicting innovations. [5,139] However, a less common but notable application is using patent analyses to guide future policy, research, and strategy, similar to the "bottom-up" approach of the European Commission’s Joint Research Centre for detecting weak signals.[140] This approach can also reveal research gaps, aiding in directing future funding and policies to address these gaps.

Not surprisingly, cancer, respiratory conditions and infections were the top three topics addressed by the included studies. Considering that this scoping review limited the included studies to only those published between 2020 and 2023 it was expected that COVID or, more broadly, respiratory conditions appeared in our dataset quite prominently. However, the focus on cancer technologies may respond to the growing burden of cancer worldwide and its impact in underserved populations.[141] Likewise the emphasis on infection topics may respond to the antimicrobial resistance crisis that we are currently facing as well as the challenges posed by the growth of communicable diseases worldwide that continue to be the cause of substantial morbidity and mortality.[142] Our review identified a wide range of technologies for which patent analyses were undertaken (Table 1, Appendix B), however we could not establish a methodological link between analyses methods and technology types. This could be due to only including studies published between 2020-2023, a longer time limit would have yielded more historical data that could have allowed some trend analysis on patent studies by technology type. Notwithstanding, our results suggest the broad applicability of patent studies to any type of technology.

The analysis showed that tools used for patent data analysis primarily focus on creating graphs and network visualizations, with examples including Gephi,[143] VOSViewer[144], and Themescape Map[145]. Data manipulation tools like Pandas (Python)[146] also played a key role. Since there is no established framework for analyzing patent data to guide horizon scanning methods, the review highlighted that data transformation and visualization are critical for detecting emerging trends. Additionally, Python and R were the most common programming languages used to develop custom tools.

## Conclusions

Our findings highlight a broad range of methodological approaches and the importance of patent data in detecting emerging trends. Rapid scoping methods proved effective, supported by precise patent-related keywords that enhanced search sensitivity. The review uncovered nearly 200 different sources for patent data retrieval, with many studies employing multiple sources, although few addressed de-duplication of data—a critical step to prevent data inflation and ensure accurate analyses of innovation clusters. Beyond trend analysis, patent data also informs policy, research, and strategy, in line with the European Commission’s "bottom-up" horizon scanning approach. Our review highlighted a focus on cancer, respiratory conditions, and infections, reflecting global health challenges and recent pandemic impacts. The use of tools for data manipulation and visualization, including Gephi, VOSViewer, and programming languages like Python and R, demonstrated the centrality of robust data transformation and visualisation for horizon scanning. Moving forward, the Innovation Observatory will engage in establishing frameworks for patent data analysis which will feed into a broader framework for underpinning future MedTech innovation identification at patent or early stage.

## Supporting information

Appendix B. Table of included studies

Appendix A. Search strategies

## Data Availability

All data produced in the present work are contained in the manuscript and supplementary materials.

