## Appendix B. Table of included studies for "MedTech innovation identification: A rapid scoping review of patent research studies to inform horizon scanning methods"

Table 1. Characteristics of included studies

| | Publication details | Type of Research Paper | Type of Technology | Type of Source |
|---|---|---|---|---|
| **1** | Hachimi A et al. A 20-year patent review and innovation trends on hydrogel-based coatings used for medical device biofabrication. 2022[14] | Review | Hydrogel-based coatings on medical devices | Patents |
| **2** | Borzova E et al. The Patent Landscape Analysis of Skin Bioinks for 3D Bioprinting. 2022[15] | Original Research | Skin bioinks | Patents |
| **3** | Tarasova EV et al. Actinomycetes as Producers of Biologically Active Terpenoids: Current Trends and Patents. 2023[16] | Review | Natural compounds for use in medicine | Various Sources |
| **4** | Asif M et al. Advancements, Trends and Future Prospects of Lower Limb Prosthesis. 2021[17] | Review | Prosthetics | Patents |
| **5** | Cammarano A et al. Advances in Transdermal Drug Delivery Systems: A Bibliometric and Patent Analysis. 2023[18] | Review | Transdermal drug delivery | Various Sources |
| **6** | Mitsumori Y et al. An Analysis of COVID-19 Related IPRs: Should they be Promoted, Waived or Pooled? 2022[19] | Review | Vaccines and pharmaceuticals | Patents |
| **7** | Ge J et al. Analysis of patent development status of lipid nanoparticle delivery system for mRNA vaccines. 2022[20] | Original Research | mRNA vaccines | Patents |

| | | | | |
|---|---|---|---|---|
| **8** | Danylenko YA et al. Analysis of scintillation materials for nuclear medicine on the basis of patent analytics. 2023[21] | Original Research | Component of medical diagnostic devices | Patents |
| **9** | Oda T et al, An analysis of the key drivers of the Japanese digital therapeutics patents: A cross-sectional study. 2023[22] | Original Research | Digital therapeutics | Patents |
| **10** | Wei F et al. Analysis of trends in patent development for coronavirus detection, prevention, and treatment technologies in key countries. 2021{Wei, 2022 #1844} | Review | Detection, vaccines, and treatment | Patents |
| **11** | Lee JH et al. Analysis of trends in patents on insect-derived medicinal materials for skin diseases. 2020[23] | Original Research | Insect-derived medicinal materials | Patents |
| **12** | Silva M et al. Antarctic organisms as a source of antimicrobial compounds: a patent review. 2022[24] | Review | Antimicrobial compounds | Patents |
| **13** | Wu NJW et al. The Application of Nanotechnology for Quantification of Circulating Tumour DNA in Liquid Biopsies: A Systematic Review. 2022[25] | Review | Nanotechnology | Various Sources |
| **14** | Ragno L et al. Application of Social Robots in Healthcare: Review on Characteristics, Requirements, Technical Solutions. 2023[26] | Review | Social robots | Various Sources |
| **15** | Borge L et al. Assessing Interdisciplinary Research Within an Emerging Technology Network: A Novel Approach Based on Patents in the Field of Bioplastics. 2022[27] | Original Research | Bioplastics | Patents |

| | | | | |
|---|---|---|---|---|
| **16** | Sertkaya A et al. Assessing the state of antibacterial drug discovery through patent analysis. 2023[28] | Short communication | Antibacterial drugs | Patents |
| **17** | Cañete P et al. Assistive Technology to Improve Collaboration in Children with ASD: State-of-the-Art and Future Challenges in the Smart Products Sector. 2022[29] | Review | Assistive technology | Various Sources |
| **18** | Liu XX et al. Bibliometric Study of Adaptogens in Dermatology: Pharmacophylogeny, Phytochemistry, and Pharmacological Mechanisms. 2023[30] | Review | Medicinal Plants | Various Sources |
| **19** | Yuan YJ et al. CAR-based cell therapy: evaluation with bibliometrics and patent analysis. 2021[31] | Review | CAR-based cell therapy | Various Sources |
| **20** | Speziali M et al. Cellulose technologies applied to biomedical purposes from the patentometric point of view. 2020[32] | Original Research | Cellulose technologies | Patents |
| **21** | Tiwari A et al. Cheminformatics: A Patentometric Analysis. 2022[33] | Original Research | Cheminformatics | Patents |
| **22** | Chuah LH et al. Chitosan based drug delivery systems for skin atopic dermatitis: recent advancements and patent trends. 2023[34] | Review | Chitosan based drug delivery systems | Patents |
| **23** | Altuntas S et al. A clustering-based approach for the evaluation of candidate emerging technologies. 2020[35] | Original Research | Dental implants | Patents |
| **24** | Lee S et al. Comparing technology convergence of artificial intelligence on the industrial sectors: two-way | Original Research | Cross-industry | Patents |

| | | | | |
|---|---|---|---|---|
| | approaches on network analysis and clustering analysis. 2021[36] | | | |
| **24** | Fluit R et al. A Comparison of Control Strategies in Commercial and Research Knee Prostheses. 2019[37] | Review | Knee prostheses | Various Sources |
| **26** | Melo RL et al. A comprehensive review on enzyme-based biosensors: Advanced analysis and emerging applications in nanomaterial-enzyme linkage. 2024[38] | Review | Enzyme based biosensors | Various Sources |
| **27** | Sanchez-Campos N, et al. Conotoxin Patenting Trends in Academia and Industry. 2022[39] | Review | Conotoxins | Patents |
| **28** | Ailia MJ, et al. Current Trend of Artificial Intelligence Patents in Digital Pathology: A Systematic Evaluation of the Patent Landscape. 2022[40] | Review | Digital pathology | Patents |
| **29** | Chen Y, et al. Delivery of therapeutic small interfering RNA: The current patent-based landscape. 2022[41] | Original Research | siRNA delivery technologies | Patents |
| **30** | Bhatnagar P, et al. Delivery systems for platelet derived growth factors in wound healing: A review of recent developments and global patent landscape. 2022[42] | Review | Delivery systems for platelet derived growth factors | Patents |
| **31** | Valadas LAR, et al. Development and innovation on dental products in Argentina: A technological prospecting based on patents. 2020[43] | Original Research | Dental products | Patents |
| **32** | Xu CM, et al. The Development of Marine Drugs: A Research Based on Patent Analysis. 2020[44] | Original Research | Marine drugs | Patents |

| | | | | |
|---|---|---|---|---|
| **33** | Imran M, et al. Development of Therapeutic and Prophylactic Zinc Compositions for Use against COVID-19: A Glimpse of the Trends, Inventions, and Patents. 2022[45] | Review | Prophylactic Zinc Compositions | Patents |
| **34** | Chang SH. The development trend and academic patent technology network of laser and optical technologies. 2021[46] | Original Research | Laser and optical technologies | Patents |
| **35** | Xin Y, et al. The development trend of artificial intelligence in medical: A patentometric analysis. 2021[47] | Original Research | Medical | Patents |
| **36** | Singh M, et al. Diagnostic and therapeutic approaches for endometriosis: a patent landscape. 2023[48] | Review | Diagnostics and therapeutics | Patents |
| **37** | Litvinova O, et al. Digital Pills with Ingestible Sensors: Patent Landscape Analysis. 2022[49] | Review | Digital pills with sensors | Various Sources |
| **38** | Imran M, et al. Discovery, Development, and Patent Trends on Molnupiravir: A Prospective Oral Treatment for COVID-19. 2021[50] | Review | Molnupiravir | Patents |
| **39** | Imran M, et al. Discovery, Development, Inventions, and Patent Trends on Mobocertinib Succinate: The First-in-Class Oral Treatment for NSCLC with EGFR Exon 20 Insertions. 2021[50] | Review | Mobocertinib Succinate | Patents |
| **40** | Jeon D, et al. A doc2vec and local outlier factor approach to measuring the novelty of patents. 2021[51] | Original Research | Medical Imaging | Patents |
| **41** | Sharma R, et al. Drug Discovery, Diagnostic, and therapeutic trends on Mpox: A patent landscape. 2021[52] | Original Research | Drug Discovery, Diagnostic, and therapeutics | Patents |

| | | | | |
|---|---|---|---|---|
| **42** | Mohajel N, et al. Ebola as a case study for the patent landscape of medical countermeasures for emerging infectious diseases. 2021[53] | Original research | Diagnostic tests and vaccines | Patents |
| **43** | Picanco-Castro V, et al. Emerging CAR T cell therapies: clinical landscape and patent technological routes. 2020[54] | Review | CAR T cell therapies | Various Sources |
| **44** | Zhou WY, et al. Emerging Patent Landscape for Gene Therapy as a Potential Cure for COVID-19. 2021[55] | Review | Gene therapy | Various Sources |
| **45** | Picanco-Castro V, et al. Emerging patent landscape for non-viral vectors used for gene therapy. 2020{Picanco-Castro, 2020 #1910} | Original research | Gene therapy | Various Sources |
| **46** | Abdi S, et al. Emerging technologies and their potential for generating new assistive technologies. 2021[56] | Original research | Assistive technology | Various Sources |
| **47** | Wadhawa R, et al. Exploring the landscape of genetics patents in the United States from 2005 to 2020. 2022[57] | Original research | Genetics | Patents |
| **48** | Robinson AA, et al. Examining the Role of Actors in an Emerging Technological System: The Case of POC Devices. 2023[58] | Original research | Micro/nanofluidic-based point-of-care (mnPOC) devices | Various Sources |
| **49** | Jeon E, et al. Exploring new digital therapeutics technologies for psychiatric disorders using BERTopic and PatentSBERTa. 2021[59] | Original research | Digital therapeutics (DTx) | Patents |
| **50** | Gadiya Y, et al. Exploring SureChEMBL from a drug discovery perspective. 2024[60] | Original research | Pharmaceutical drugs | Patents |

| | | | | |
|---|---|---|---|---|
| **51** | Wang YH. Exploring Technology-Driven Technology Roadmaps (TRM) for Wearable Biosensors in Healthcare. 2024[61] | Original research | Wearable biosensors | Various Sources |
| **52** | Singh P, et al. Ficus benghalensis-A comprehensive review on pharmacological research, nanotechnological applications, and patents. 2023[62] | Review | Ficus benghalensis | Various Sources |
| **53** | Culmone C, et al. Follow-The-Leader Mechanisms in Medical Devices: A Review on Scientific and Patent Literature. 2021[63] | Review | Medical Devices | Various Sources |
| **54** | Zagoya-Lopez Z, et al. Foot/Ankle Prostheses Design Approach Based on Scientometric and Patentometric Analyses. 2021[64] | Review | Foot/Ankle protheses | Various Sources |
| **55** | Lyu L, et al. The global chimeric antigen receptor T (CAR-T) cell therapy patent landscape. 2020[65] | Original research | CAR-T cell therapy | Patents |
| **56** | Frisio DG, et al. Global Innovation Trends for Plant-Based Vaccines Production: A Patent Analysis. 2021[66] | Original research | Plant-based vaccines | Patents |
| **57** | Liu K, et al. Global landscape of patents related to human coronaviruses. 2021[67] | Review | Human Coronaviruses | Patents |
| **58** | Li M, et al. The global mRNA vaccine patent landscape. 2022{Li, 2022 #1931} | Original research | RNA vaccines | Patents |
| **59** | Braga L, et al. The global patent landscape of artificial intelligence applications for cancer. 2023[68] | Original research | AI cancer applications | Patents |

| | | | | |
|---|---|---|---|---|
| **69** | Liu K, et al. Global Patent Landscape of Benign Prostatic Hyperplasia Drugs. 2022[69] | Original research | Benign prostatic hyperplasia (BPH) drugs | Patents |
| **61** | Cai Y, et al. The global patent landscape of emerging infectious disease monkeypox. 2024[70] | Original research | Monkeypox | Patents |
| **62** | Li Q, et al. The global patent landscape of HER2-targeted biologics. 2023[71] | Original research | HER2-targeted therapies | Patents |
| **63** | Lyu M, et al. The global patent landscape of mRNA for diagnosis and therapy. 2023[72] | Original research | Messenger RNA (mRNA) | Patents |
| **64** | Liu K, et al. Global research on artemisinin and its derivatives: Perspectives from patents. 2020[73] | Review | Artemisinin derivatives | Patents |
| **65** | Maresova P, et al. Health–Related ICT Solutions of Smart Environments for Elderly–Systematic Review. 2020[74] | Review | ICT for smart environments | Various Sources |
| **66** | Zhou W, et al. Human gene therapy: A patent analysis. 2021[75] | Review | Gene therapy | Patents |
| **67** | Machuca-Martinez F, et al. Coronaviruses: A patent dataset report for research and development (R&D) analysis. 2020[76] | Original research | Coronaviruses | Patents |
| **68** | Shin HJ, et al. Identifying Areas of Technology Commercialization in the Biomedical Sector: An Integrated Analysis of Patents and Publications. 2022[77] | Original research | Biomedical technologies | Other |
| **69** | Raghu Kiran, CVS, et al. Idiom of gastroretentive drug delivery systems: Comprehensive view on innovation technologies, patents and clinical [trails]. 2023[78] | Review | Gastroretentive drug delivery | Various Sources |

| | | | | |
|---|---|---|---|---|
| **70** | Imran M, et al. Innovations and patent trends in the development of USFDA approved protein Kinase inhibitors in the last two decades. 2021[79] | Review | Protein Kinase Inhibitors | Patents |
| **71** | Aboy M, et al. Mapping the European patent landscape for medical uses of known products. 2021[80] | Original Research | N/A | Patents |
| **72** | Aboy M, et al. Mapping the patent landscape of medical machine learning. 2023[81] | Original Research | Machine Learning | Patents |
| **73** | Burgio V, et al. Mechanical Stapling Devices for Soft Tissue Repair: A Review of Commercially Available Linear, Linear Cutting, and Circular Staplers. 2024[82] | Review | Mechanical Stapling Devices | Various Sources |
| **74** | Queiroz AAFLN, et al. mHealth Strategies Related to HIV Postexposure Prophylaxis Knowledge and Access: Systematic Literature Review, Technology Prospecting of Patent Databases, and Systematic Search on App Stores. 2021[83] | Review | Mobile health (mHealth) interventions | Various Sources |
| **75** | Lohita S, et al. Myocardial Infarction: Background, Recent Advances, and Interventions Supported by Clinical Trial and Patent Landscape. 2023[84] | Review | Not specified | Various Sources |
| **76** | Zhang HL, et al. New Frontier in Antiviral Drugs for Disorders of the Respiratory System. 2022[85] | Original research | Antiviral drugs | Patents |
| **77** | Mancilla-de-la-Cruz J, et al. The Next Pharmaceutical Path: Determining Technology Evolution in Drug | Original research | Additive | Various Sources |

| | | | | |
|---|---|---|---|---|
| | Delivery Products Fabricated with Additive Manufacturing. 2020[86] | | | |
| **78** | Riondato M, et al. Oldie but Goodie: Is Technetium-99m Still a Treasure Trove of Innovation for Medicine? A Patents Analysis (2000−2022). 2023[87] | Original research | Technetium-99m | Patents |
| **79** | Colonia BSO, et al. Omega-3 microbial oils from marine thraustochytrids as a sustainable and technological solution: A review and patent landscape. 2020[88] | Original research | Omega-3 microbial oils | Various Sources |
| **80** | Ma J, et al. Organization oriented technology opportunities analysis based on predicting patent networks: a case of Alzheimer's disease. 2022[89] | Original research | Technologies (not specified) | Patents |
| **81** | Litvinova O, et al. Patent analysis of digital sensors for continuous glucose monitoring. 2023[90] | Review | Digital sensors | Various Sources |
| **82** | Klongthong W, et al. A Patent Analysis to Identify Emergent Topics and Convergence Fields: A Case Study of Chitosan. 2021[91] | Original research | Chitosan | Patents |
| **83** | Hani U, et al. Patent bibliometrics in spinal deformity: the first bibliometric analysis of spinal deformity's technological literature. 2023[92] | Original research | Surgical devices | Patents |
| **84** | Devarapalli P, et al. Patent intelligence of RNA viruses: Implications for combating emerging and re-emerging RNA virus based infectious diseases. 2022[93] | Original research | RNA viruses | Patents |
| **85** | Hernández-Melchor D, et al. The patent landscape in the field of stem | Original research | Stem cell therapy | Patents |

| | | | | |
|---|---|---|---|---|
| | cell therapy: closing the gap between research and clinic. 2024[94] | | | |
| **86** | Greenberg A, et al. Patent landscape of brain–machine interface technology. 2021[95] | Original research | Brain–machine interface | Patents |
| **87** | Cho YD, et al. Patent landscape report on dental implants: A technical analysis. 2021[96] | Original research | Dental implants | Patents |
| **88** | Litvinova O, et al. Patent landscape review of non-invasive medical sensors for continuous monitoring of blood pressure and their validation in critical care practice. 2023[97] | Original research | Non-invasive medical sensors for continuous monitoring of blood pressure | Various Sources |
| **89** | Francis N, et al. Patent Landscape Review on Ankle Sprain Prevention Method: Technology Updates. 2023[98] | Original research | Ankle Sprain Prevention technology | Patents |
| **90** | Juiz PJ, et al. Patent Mining on the Use of Antioxidant Phytochemicals in the Technological Development for the Prevention and Treatment of Periodontitis. 2024[99] | Original research | Antioxidant Phytochemicals | Patents |
| **91** | Chartoumpekis DV, et al. Patent Review (2017-2020) of the Keap1/Nrf2 Pathway Using PatSeer Pro: Focus on Autoimmune Diseases. 2020[100] | Original research | Nuclear factor erythroid 2-related factor 2 (Nrf2) and cytoplasmic inhibitor Kelch-like ECH-associated protein 1 (Keap1 | Patents |
| **92** | Parihar K, et al. A patent review on strategies for biological control of mosquito vector. 2020[101] | Original research | Biological control of mosquito | Patents |
| **93** | Xiong YH, et al. Patented technologies for schistosomiasis | Original research | Medicines, devices | Patents |

| | | | | |
|---|---|---|---|---|
| | control and prevention filed by Chinese applicants. 2021[102] | | | |
| **94** | Russo Serafini M, et al. The Patenting and Technological Trends in Hernia Mesh Implants. 2020[103] | Review | Prosthetic surgical meshes | Various Sources |
| **95** | Mendez CRA, et al. Patentometric analysis of the technological development of Biotechnology for health in higher education institutions in Rio Grande do Sul. 2024[104] | Review | Biotechnology in healthcare | Patents |
| **96** | Gkika DA, et al. Patents of nanomaterials related with cancer treatment applications. 2020[105] | Review | Nanomaterials related with cancer treatment applications | Patents |
| **97** | Gadiya Y, et al. Pharmaceutical patent landscaping: A novel approach to understand patents from the drug discovery perspective. 2023[106] | Review | Pharmaceuticals and biotechnology | Patents |
| **98** | Patel S, et al. Probiotic Formulations: A Patent Landscaping Using the Text Mining Approach. 2022[107] | Review | Probiotics | Patents |
| **99** | Shivakumar P, et al. Prospection of chitosan and its derivatives in wound healing: Proof of patent analysis. 2021[108] | Review | Chitosan | Various Sources |
| **100** | Kurakula M, et al. Prospection of recent chitosan biomedical trends: Evidence from patent analysis. 2020[109] | Review | Chitosan | Various Sources |
| **101** | Islam MM, et al. The Race to Replace PDE5i: Recent Advances and Interventions to Treat or Manage Erectile Dysfunction: Evidence from | Review | Phosphodiesterase type 5 inhibitor | Various Sources |

| | | | | |
|---|---|---|---|---|
| | Patent Landscape (2016–2021). 2022[110] | | | |
| **102** | Durmuşoğlu A, et al. Remembering Medical Ventilators and Masks in the Days of COVID-19: Patenting in the Last Decade in Respiratory Technologies. 2022[111] | Original research | Medical Ventilators and Masks | Patents |
| **103** | Zhang T, et al. The research activities and development trends of antineoplastics targeting PD-1/PD-L1 based on scientometrics and patentometrics. 2022[112] | Conference paper | Antineoplastics | Patents |
| **104** | DasNandy A, et al. A review of patent literature on the regulation of glucose metabolism by six phytocompounds in the management of diabetes mellitus and its complications. 2023[113] | Review | Phytocompounds | Patents |
| **105** | Yeh TF, et al. A review of technological developments in lipid nanoparticle application for mRNA vaccination. 2023[114] | Original research | Lipid nanoparticle | Various Sources |
| **106** | Wang Q, et al. A Scientometric Analysis and Visualization of Scientific Research and Technology Innovation in Needle-free Insulin Injection From 1974 to 2022. 2023[115] | Original research | Needle-free insulin injection | Various Sources |
| **107** | Jiang J, et al. The state of the art and future trends of root canal files from the perspective of patent analysis: a study design. 2022[116] | Review | Root canal files | Patents |
| **108** | Kong X, et al. STING as an emerging therapeutic target for drug discovery: | Review | Stimulator of interferon genes (STING | Patents |

| | | | | |
|---|---|---|---|---|
| | Perspectives from the global patent landscape. 2022[117] | | | |
| **109** | Hazis NUA, et al. Systematic Patent Review of Nanoparticles in Drug Delivery and Cancer Therapy in the Last Decade. 2021[118] | Review | Nanoparticles | Patents |
| **110** | Verma R. A Technical Analysis of MIOT in Sensitive Aspect. 2023[119] | Conference paper | Biosensors | Various Sources |
| **111** | Barragán-Ocaña A, et al. Technological development and patent analysis: the case of biopharmacy in the world and in Latin America. 2022[120] | Original research | Biopharmaceuticals | Patents |
| **112** | Hwang J, et al. Technological Opportunity Analysis: Assistive Technology for Blind and Visually Impaired People. 2020[121] | Original research | Visual assistive device | Patents |
| **113** | Liu J, et al. Technology Forecasting based on Topic Analysis and Social Network Analysis: A Case Study Focusing on Gene Editing Patents. 2021[122] | Original research | Gene editing technology | Patents |
| **114** | Wadhwa RR, et al. Temporal Trends in the United States Patent Landscape:<br>Innovation in Cardiology Across Industry and Academia. 2023[123] | Original research | Diagnostics and therapeutics in medical care | Patents |
| **115** | Erzurumlu SS, et al. Topic modeling and technology forecasting for assessing the commercial viability of healthcare innovations. 2020[124] | Original research | Healthcare innovations | Patents |
| **116** | Pasek JE, et al. Trends in bioengineering patents granted 2000-2019. 2021[125] | Original research | Bioengineering | Patents |

| | | | | |
|---|---|---|---|---|
| **117** | Chowdhury AR, et al. The trends in CRISPR research: A patent and literature study with a focus on India. 2021[126] | Review | Clustered regularly interspaced short palindromic repeat (CRISPR) | Various Sources |
| **118** | Almeida FLC, et al. Erratum to "Trends in lipase immobilization: Bibliometric review and patent analysis". 2021[127] | Review | Biotechnology | Various Sources |
| **119** | Bacigalupo ML, et al. Unveiling patenting strategies of therapeutics and vaccines: evergreening in the context of COVID-19 pandemic. 2023[128] | Original research | Therapeutics and vaccines | Patents |
| **120** | Chen TA, et al. Using Big Data Analytics on Health Industry Development: The Empirical Intellectual Property Analysis from Stem Cell Therapy. 2021[129] | Original research | Stem cell therapy | Patents |
| **121** | Liu K, et al. What, Where When and How of COVID-19 Patents Landscape: A Bibliometrics Review. 2022[130] | Original research | COVID-19 related technologies | Patents |
| **122** | Rincon-Lopez J, et al. When Cyclodextrins Met Data Science: Unveiling Their Pharmaceutical Applications through Network Science and Text-Mining. 2021[131] | Original research | Cyclodextrins | Patents |
| **123** | Kim WJ, et al. The worldwide patent landscape of dental implant technology. 2022[132] | Review | Dental implants | Patents |
| **124** | Azman AA, et al. Worldwide trend discovery of structural and functional relationship of metallo-β-lactamase for structure-based drug design: A | Review | Metallo-β-lactamase | Various Sources |

| | |
|---|---|
| | bibliometric evaluation and patent analysis. 2023[133] |
