## Appendix A. Search strategies for "MedTech innovation identification: A rapid scoping review of patent research studies to inform horizon scanning methods"

Full search strategies used for the identification of patent research studies

**Database:** Embase <1996 to 2024 Week 22>
**URL:** Ovid via University library link
**Data of search:** 06/06/2024
**Number of retrieved records:** 331
**Search strategy:**

| | | |
|---|---|---|
| 1 | patent* research.ti,ab,kf,kw. | 59 |
| 2 | patent* analys*.ti,ab,kf,kw. | 317 |
| 3 | patent* landscape.ti,ab,kf,kw. | 253 |
| 4 | (patent* adj3 trend*).ti,ab,kf,kw. | 241 |
| 5 | (patent* adj3 mining).ti,ab,kf,kw. | 42 |
| 6 | or/1-5 | 812 |
| 7 | limit 6 to yr="2020 -Current" | 331 |

**Notes:**
Downloaded into Endnote for de-duplication.
**Database: IEEE Xplore Digital library**
**URL: via University library link**
**Data of search: 06/06/2024**
**Number of retrieved records: 580**
**Search strategy:**
("Publication Title": patent research OR "Abstract": patent research OR "Author Keywords": patent research OR "Document Title": patent analysis OR "Abstract": patent analysis OR "Author Keywords": patent analysis OR "Document Title": patent analyses OR "Abstract": patent analyses OR "Author Keywords": patent analyses OR "Publication Title": patent landscape OR "Abstract": patent landscape OR "Author Keywords": patent landscape OR "Publication Title": patent trend OR "Abstract": patent trend OR "Author Keywords": patent trend OR "Publication Title": patent trends OR "Abstract": patent trends OR "Author Keywords": patent trends OR "Publication Title": patent mining OR "Abstract": patent mining OR "Author Keywords": patent mining)
Publication types filter: Conferences, Journals, Early Access Articles
Time limits applied: 2020 - 2024
**Notes:**
All downloaded in csv format.

**Database:** Web of Science
Institute for Scientific Information (2000) Web of science. Philadelphia, PA]: Thomson Reuters.
**URL:** via University link
**Date of search:** 06/06/2024
**Number of retrieved records:** 830
**Search strategy:**
(TS=("patent research") OR TI=("patent research") OR AB=("patent research")) OR (TS=("patent analysis") OR TI=("patent analysis") OR AB=("patent analysis")) OR (TS=("patent analyses") OR TI=("patent analyses") OR AB=("patent analyses")) OR (TS=("patent landscape") OR TI=("patent landscape") OR AB=("patent landscape")) OR (TS=("patent trend") OR TI=("patent trend") OR AB=("patent trend")) OR (TS=("patent trends") OR TI=("patent trends") OR AB=("patent trends")) OR (TS=("patent mining") OR TI=("patent mining") OR AB=("patent mining"))
Publication date limits: 2020, 2021, 2022, 2023
**Notes:**
All exported into Endnote
